# A Time Series Analysis of Trends in Medicare Utilization and Reimbursement for Cancer Immunotherapy Drugs: 2012-2017

**DOI:** 10.1101/2020.06.27.20141721

**Authors:** Pranav Puri, Denis Cortese, Sujith Baliga

## Abstract

The adoption of immunotherapy has dramatically transformed the landscape of oncology, and improved the prognosis of patients diagnosed with metastatic malignancies. Fee-for-service (FFS) Medicare is the largest payer in the U.S, and pays for physician-administered oncolytics through its medical Part B benefit. However, to the best of our knowledge, trends in Medicare utilization and expenditures for cancer immunotherapy have yet to be described. The aim of this study was to describe trends in Medicare immunotherapy reimbursement. We utilized the public Drugs@FDA database to identify all immunotherapy drugs that were approved by the FDA for solid and hematological cancers in adults as of December 31, 2017. The Medicare Physician Supplier and Other Provider Public Use File (POSPUF) provides reimbursement and utilization data on all services and procedures provided to Medicare fee-for-service beneficiaries. Using the Medicare POSPUF, we aggregated the volume of services, average Medicare reimbursement, and the number of distinct patients receiving each immunotherapy drug from January 2012 to December 2017. The number of Medicare reimbursed cancer immunotherapy drugs increased from 3 in 2012 to 12 in 2017. Over this time period, the number of patients receiving immunotherapy administrations increased 88% from 59,506 to 111,577. Similarly, the number of drug administrations increased by 2,412% from 2,338,691 to 58,752,804. From 2012 to 2017, total Medicare expenditure on cancer immunotherapy drugs increased 154% from $771,434,031 to $1,962,279,164. Medicare expenditure on cancer immunotherapy drugs accounted for 26% of the increase in total Medicare Part B drug expenditures over this time period.

## Introduction

The adoption of immunotherapy has dramatically transformed the landscape of oncology, and improved the prognosis of patients diagnosed with metastatic malignancies.^1^ Over the last five years, a number of these drugs have received US Food and Drug Administration (FDA) market authorization. In addition, the treatment of patients with immunotherapy represents a growing component of outpatient oncology practice. Oncology practices are reimbursed through a “buy and bill” system in which practices purchase drugs and bill insurers for their administration. Fee-for-service (FFS) Medicare is the largest payer in the U.S, and pays for physician-administered oncolytics through its medical Part B benefit.^2^ However, to the best of our knowledge, trends in Medicare utilization and expenditures for cancer immunotherapy have yet to be described. The aim of this study was to describe trends in Medicare immunotherapy reimbursement.

## Methods

We utilized the public Drugs@FDA database to identify all immunotherapy drugs that were approved by the FDA for solid and hematological cancers in adults as of December 31, 2017.^3^

The Medicare Physician Supplier and Other Provider Public Use File (POSPUF) provides reimbursement and utilization data on all services and procedures provided to Medicare fee-for-service beneficiaries.^4^ Using the Medicare POSPUF, we aggregated the volume of services, average Medicare reimbursement, and the number of distinct patients receiving each immunotherapy drug from January 2012 to December 2017. We restricted our analysis to claims listing physician offices as the site of service. For each drug, we estimated total Medicare expenditure by multiplying the average Medicare reimbursement by the volume of services. We also calculated aggregate annual expenditure on cancer immunotherapy as a fraction of overall Medicare Part B drug expenditure.

This study was exempt from IRB review as the dataset is publicly available and contains no patient identifiers. Data analysis was conducted using JMP version 14.

## Results

The number of Medicare reimbursed cancer immunotherapy drugs increased from 3 in 2012 to 12 in 2017. Over this time period, the number of patients receiving immunotherapy administrations increased 88% from 59,506 to 111,577. Similarly, the number of drug administrations increased by 2,412% from 2,338,691 to 58,752,804.

From 2012 to 2017, total Medicare expenditure on cancer immunotherapy drugs increased 154% from $771,434,031 to $1,962,279,164 (Table 1). During the same time period, overall Medicare Part B drug expenditure increased 49% from $9,234,672,715 to $13,761,206,920. Medicare expenditure on cancer immunotherapy drugs accounted for 26% of the increase in total Medicare Part B drug expenditures over this time period. Expenditure on cancer immunotherapy drugs comprised 8% of total Medicare Part B drug spending in 2012 and increased to 14% in 2017 (Figure 1).

**Table 1:**
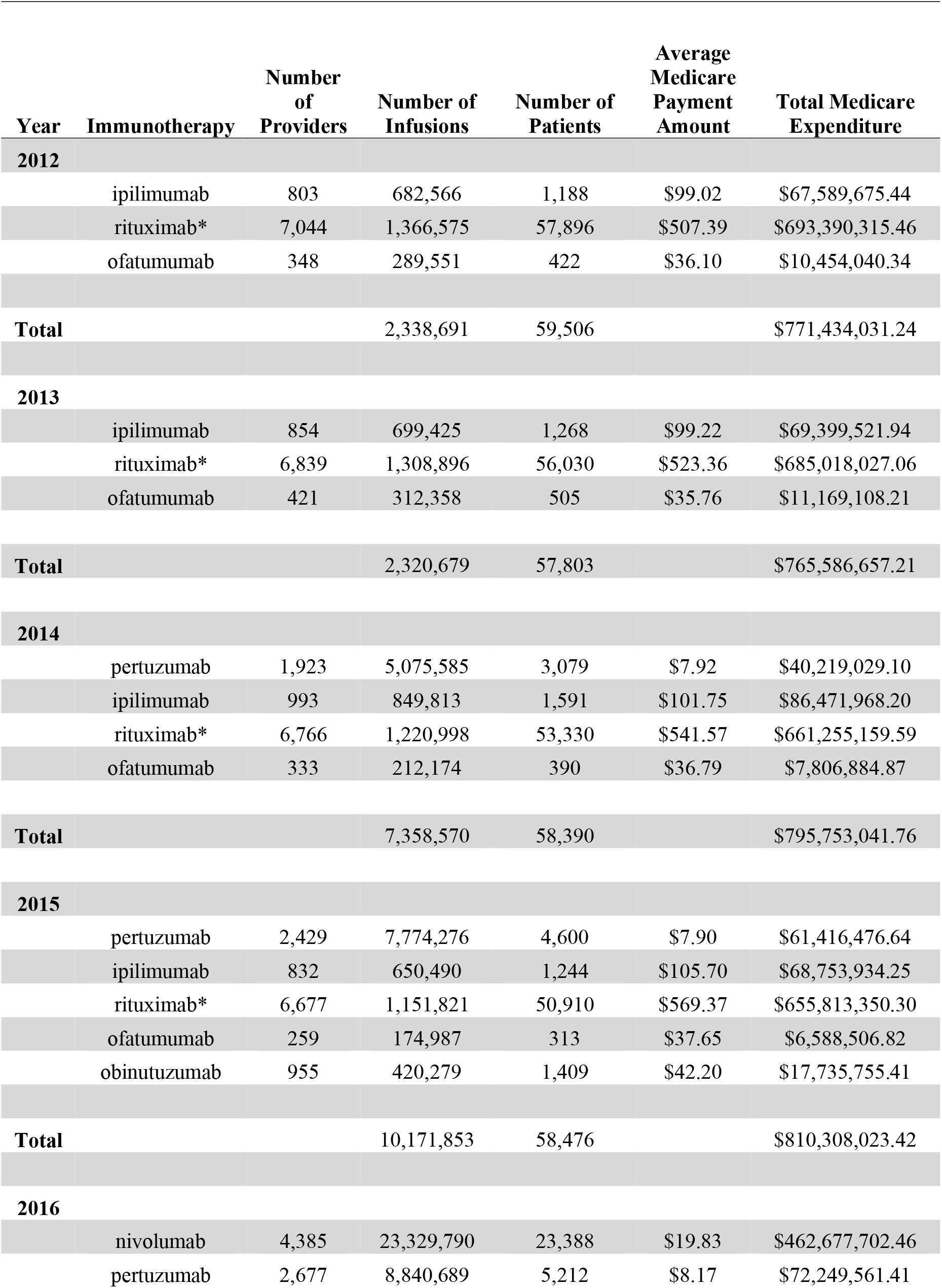
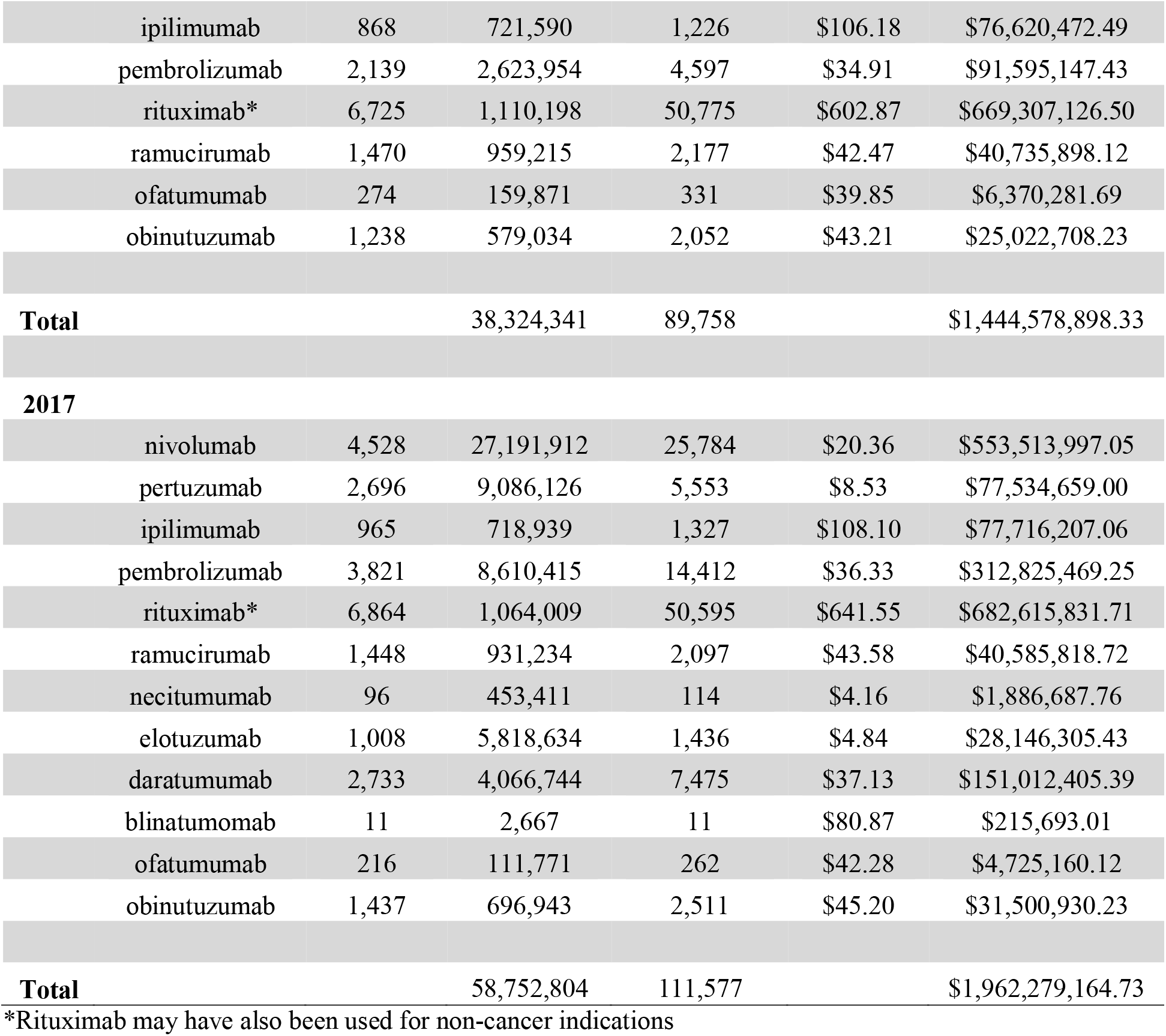
Trends in Utilization and Reimbursement of Cancer Immunotherapy Drugs

| Year | Immunotherapy | Number of Providers | Number of Infusions | Number of Patients | Average Medicare Payment Amount | Total Medicare Expenditure |
| --- | --- | --- | --- | --- | --- | --- |
| **2012** |  |  |  |  |  |  |
| | ipilimumab | 803 | 682,566 | 1,188 | \$99.02 | \$67,589,675.44 |
| | rituximab* | 7,044 | 1,366,575 | 57,896 | \$507.39 | \$693,390,315.46 |
| | ofatumumab | 348 | 289,551 | 422 | \$36.10 | \$10,454,040.34 |
| **Total** | | | 2,338,691 | 59,506 | | \$771,434,031.24 |
| **2013** |  |  |  |  |  |  |
| | ipilimumab | 854 | 699,425 | 1,268 | \$99.22 | \$69,399,521.94 |
| | rituximab* | 6,839 | 1,308,896 | 56,030 | \$523.36 | \$685,018,027.06 |
| | ofatumumab | 421 | 312,358 | 505 | \$35.76 | \$11,169,108.21 |
| **Total** | | | 2,320,679 | 57,803 | | \$765,586,657.21 |
| **2014** |  |  |  |  |  |  |
| | pertuzumab | 1,923 | 5,075,585 | 3,079 | \$7.92 | \$40,219,029.10 |
| | ipilimumab | 993 | 849,813 | 1,591 | \$101.75 | \$86,471,968.20 |
| | rituximab* | 6,766 | 1,220,998 | 53,330 | \$541.57 | \$661,255,159.59 |
| | ofatumumab | 333 | 212,174 | 390 | \$36.79 | \$7,806,884.87 |
| **Total** | | | 7,358,570 | 58,390 | | \$795,753,041.76 |
| **2015** |  |  |  |  |  |  |
| | pertuzumab | 2,429 | 7,774,276 | 4,600 | \$7.90 | \$61,416,476.64 |
| | ipilimumab | 832 | 650,490 | 1,244 | \$105.70 | \$68,753,934.25 |
| | rituximab* | 6,677 | 1,151,821 | 50,910 | \$569.37 | \$655,813,350.30 |
| | ofatumumab | 259 | 174,987 | 313 | \$37.65 | \$6,588,506.82 |
| | obinutuzumab | 955 | 420,279 | 1,409 | \$42.20 | \$17,735,755.41 |
| **Total** | | | 10,171,853 | 58,476 | | \$810,308,023.42 |
| **2016** |  |  |  |  |  |  |
| | nivolumab | 4,385 | 23,329,790 | 23,388 | \$19.83 | \$462,677,702.46 |
| | pertuzumab | 2,677 | 8,840,689 | 5,212 | \$8.17 | \$72,249,561.41 |

|  |  |  |  |  |  |
| --- | --- | --- | --- | --- | --- |
| ipilimumab | 868 | 721,590 | 1,226 | \$106.18 | \$76,620,472.49 |
| pembrolizumab | 2,139 | 2,623,954 | 4,597 | \$34.91 | \$91,595,147.43 |
| rituximab* | 6,725 | 1,110,198 | 50,775 | \$602.87 | \$669,307,126.50 |
| ramucirumab | 1,470 | 959,215 | 2,177 | \$42.47 | \$40,735,898.12 |
| ofatumumab | 274 | 159,871 | 331 | \$39.85 | \$6,370,281.69 |
| obinutuzumab | 1,238 | 579,034 | 2,052 | \$43.21 | \$25,022,708.23 |
| **Total** | | 38,324,341 | 89,758 | | \$1,444,578,898.33 |
| **2017** |  |  |  |  |  |
| nivolumab | 4,528 | 27,191,912 | 25,784 | \$20.36 | \$553,513,997.05 |
| pertuzumab | 2,696 | 9,086,126 | 5,553 | \$8.53 | \$77,534,659.00 |
| ipilimumab | 965 | 718,939 | 1,327 | \$108.10 | \$77,716,207.06 |
| pembrolizumab | 3,821 | 8,610,415 | 14,412 | \$36.33 | \$312,825,469.25 |
| rituximab* | 6,864 | 1,064,009 | 50,595 | \$641.55 | \$682,615,831.71 |
| ramucirumab | 1,448 | 931,234 | 2,097 | \$43.58 | \$40,585,818.72 |
| necitumumab | 96 | 453,411 | 114 | \$4.16 | \$1,886,687.76 |
| elotuzumab | 1,008 | 5,818,634 | 1,436 | \$4.84 | \$28,146,305.43 |
| daratumumab | 2,733 | 4,066,744 | 7,475 | \$37.13 | \$151,012,405.39 |
| blinatumomab | 11 | 2,667 | 11 | \$80.87 | \$215,693.01 |
| ofatumumab | 216 | 111,771 | 262 | \$42.28 | \$4,725,160.12 |
| obinutuzumab | 1,437 | 696,943 | 2,511 | \$45.20 | \$31,500,930.23 |
| **Total** | | 58,752,804 | 111,577 | | \$1,962,279,164.73 |

\*Rituximab may have also been used for non-cancer indications

**Figure 1.**
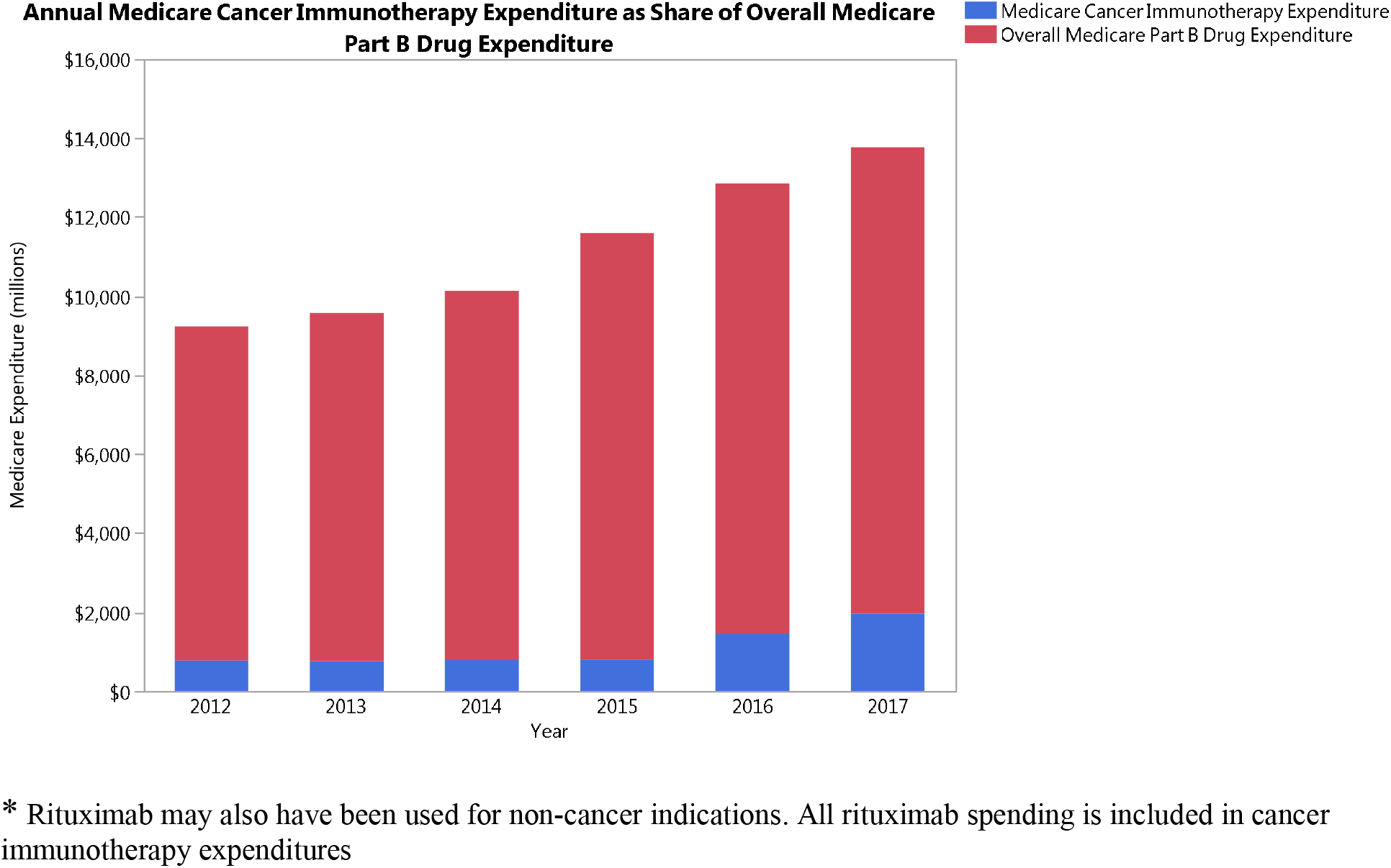
* Rituximab may also have been used for non-cancer indications. All rituximab spending is included in cancer immunotherapy expenditures.

## Discussion

As numerous immunotherapy drugs have obtained FDA approval, there has been a concomitant increase in the number of patients receiving immunotherapy. Despite encouraging results from clinical trials, only a small fraction of patients sustain response to treatment. Nevertheless, utilization has significantly increased total Medicare expenditure on cancer immunotherapy drugs. In addition, cancer immunotherapies represent a growing share of overall Medicare Part B drug spending. Prior studies have raised concerns that Medicare’s existing oncolytic reimbursement models create financial risks for independent oncology practices, while incentivizing increased utilization.^5^ Therefore, policy makers should develop value-based reimbursement models that link payments for immunotherapy use to improved patient outcomes, in particular survival and quality of life. In order to so, future studies must define the subsets of patients most likely to respond to immunotherapies. This can be achieved through emerging biomarker approaches, or through adaptive trial designs, such as those being studied in the I-SPY trials.^6^

## Data Availability

Data available on request

https://www.cms.gov/Research-Statistics-Data-and-Systems/Statistics-Trends-and-Reports/Medicare-Provider-Charge-Data/Physician-and-Other-Supplier

## Acknowledgment

Data access and responsibility: Pranav Puri had full access to all the data in the study and take responsibility for the integrity of the data and the accuracy of the data analysis. Pranav Puri and Sujith Baliga take responsibility for the integrity of the work as a whole, from inception to published article.

## References

1. Duan J, Cui L, Zhao X, et al. Use of Immunotherapy With Programmed Cell Death 1 vs Programmed Cell Death Ligand 1 Inhibitors in Patients With Cancer: A Systematic Review and Meta-analysis. JAMA Oncol. 2019.

2. Polite B, Conti RM, Ward JC. Reform of the Buy-and-Bill System for Outpatient Chemotherapy Care Is Inevitable: Perspectives from an Economist, a Realpolitik, and an Oncologist. Am Soc Clin Oncol Educ Book. 2015:e75–80.

3. United States Food and Drug Administration. Drugs@FDA: FDA-approved drugs. In:2020.

4. Medicare Provider Utilization and Payment Data: Physician and Other Supplier. 2012-2017. https://www.cms.gov/Research-Statistics-Data-and-Systems/Statistics-Trends-and-Reports/Medicare-Provider-Charge-Data/Physician-and-Other-Supplier. accessed March 08, 2020.

5. Malin JL, Weeks JC, Potosky AL, Hornbrook MC, Keating NL. Medical oncologists’ perceptions of financial incentives in cancer care. J Clin Oncol. 2013;31(5):530–535.

6. Das S, Lo AW. Re-inventing drug development: A case study of the I-SPY 2 breast cancer clinical trials program. Contemp Clin Trials. 2017;62:168–174.

